# Risk Perceptions and Private Protective Behaviors: Evidence from COVID-19 Pandemic^*^

**DOI:** 10.1101/2022.03.08.22272111

**Authors:** M. Kate Bundorf, Jill DeMatteis, Grant Miller, Maria Polyakova, Jialu L. Streeter, Jonathan Wivagg

## Abstract

We analyze data from a survey we administered during the COVID-19 pandemic to investigate the relationship between people’s subjective beliefs about risks and their private protective behaviors. On average, people substantially overestimate the absolute level of risk associated with economic activity, but have correct signals about their relative risk. Subjective risk beliefs are predictive of changes in economic activities independently of government policies. Government mandates restricting economic behavior, in turn, attenuate the relationship between subjective risk beliefs and protective behaviors.

## 1 Introduction

Subjective perceptions of risk guide decision-making in nearly all economic domains (Hurd 2009). While a rich theoretical literature highlights the importance of subjective beliefs for individual actions, fewer studies provide empirical evidence of the link between risk perceptions and behaviors, particularly in non-laboratory settings and in high-income countries (Peltzman 1975; Delavande 2014; Delavande and Kohler 2015; Mueller et al. 2021). Yet, knowing how subjective risk perceptions influence behavior is essential for policy development. Many policies, such as government insurance mandates and public health campaigns, seek to change behavior in the presence of risk. Predicting responses to such policies requires understanding how policies influence perceived risks and, in turn, how risk beliefs influence behaviors (Manski 2004).

We examine the relationship between individual perceptions of (a very salient) risk and economic behaviors, as well as how much people believe that policy interventions change this relationship. Our analysis is based on a nationally-representative survey that we fielded in the United States in May 2020, near the beginning of the COVID-19 pandemic. We collected information on individual beliefs about COVID-19 risks, including the likelihood of infection and the likely health implications of infection; how much people had changed their activities in response to the pandemic; and how much they believed that they would have changed their activities in a hypothetical scenario without government restrictions on activities.^1^

We report three main findings. First, most people substantially overestimated the absolute level of risk. On average, participants reported that their risk of contracting COVID-19 while performing an economic activity early in the pandemic was 40% to 62%, depending on the activity. The actual prevalence of COVID-19 at the time, however, was much lower. By May 31, 2020, the U.S had reported 1,786,683 cumulative cases or a prevalence of 0.5%.^2^ This is consistent with research demonstrating that, although elicited probabilistic expectations generally follow basic properties of probabilities and predict actual future decisions and behavior (e.g. Finkelstein and McGarry 2006; Dominitz and Manski 2007; Hurd 2009; Delavande et al. 2011; Hendren 2013), people also often overestimate both small and highly publicized risks (Viscusi et al. 1998). At the same time, people had more accurate perceptions about their relative risks. Individual assessment of risk varied substantially across socio-economic and demographic groups as well as across geographies in ways that were generally consistent with observational data on the variation in COVID-19 prevalence and outcomes.^3^

Second, we find that individual beliefs about risks were related to individual protective behavior. In the early stages of the COVID-19 pandemic, minimizing interpersonal interactions through activity reduction was arguably the most effective protective behavior available to reduce infection risk. Our results indicate that those who believed that they had a higher risk of infection also believed that, in the absence of policies restricting economic activity, they would privately choose to reduce their activities more in response to the pandemic across a variety of domains.^4^ This result builds on work in economic epidemiology which emphasizes the endogeneity of private protective behavior with respect to disease prevalence (Philipson 2000). Relating COVID-19 prevalence and behaviors directly, we find prevalence elasticities (a measure of responsiveness of individual protective behaviors to disease outbreak) ranging from 0.02 to 0.77 across different domains of avoidance behavior and policy environments.

Finally, our data suggest that public mandates for activity restriction attenuate the relationship between subjective risk perceptions and behavior. As intended, by restricting population-level activity, mandates reduce the externality that lower risk individuals (who, as we find, would privately choose relatively less protective behavior) exert on higher risk individuals (McAdams 2021; Adda 2016).^5^ At the same time, we also find that people who believe that they are at higher risk of an adverse outcome, conditional on infection, are relatively *more likely* to engage in economic activity in the presence of a government SIP order – consistent with the idea that policies reducing risk can lead to compensatory (or off-setting) increases in risky behavior (Ehrlich and Becker 1972; Shavell 1979).

Narrowly, our paper contributes to the rapidly growing literature on the COVID-19 pandemic, and especially the role of private beliefs and government policies in driving behaviors. Studies aimed at measuring the effect of government policies have used either microsimulation models (Davies et al. 2020; Ferguson et al. 2020; Jarvis et al. 2020; Ngonghala et al. 2020; Peak et al. 2020; Prem et al. 2020) or, increasingly, retrospective analyses of policy implementation (Chen et al. 2020; Abouk and Heydari 2021; Nguyen et al. 2020; Dave et al. 2021; Chudik et al. 2020; Glaeser et al. 2021; Gupta et al. 2020; Flaxman et al. 2020; Alexander and Karger 2020; Klein, LaRocky, et al. 2020; Klein, LaRock, et al. 2020; Jacobsen and Jacobsen 2020; Weill et al. 2020; Atkeson et al. 2020). Several contemporaneous studies have examined risk perceptions during the pandemic, including the relationship between risk perceptions and economic anxieties (Alsan et al. 2020; Bordalo et al. 2020; Wise et al. 2020; Barrios and Hochberg 2020; Fan et al. 2020; Dryhurst et al. 2020; Galasso et al. 2020; Nino et al. 2021; Reiter and Katz 2021; Akesson et al. 2020; Fetzer et al. 2021). Most studies of SIP policies, however, have placed less emphasis on subjective risk perceptions and their relationship with individual, private decisions regarding risk protection.

While the empirical patterns that we document are specific to the COVID-19 context, our findings contribute to the literature on eliciting subjective risk beliefs to understand how people make decisions under risk more broadly (Manski 2004; Hurd 2009; Delavande 2014; Delavande and Kohler 2015; Spinnewijn 2013; Handel and Kolstad 2015; Miller et al. 2020; Mueller et al. 2021). Further, our results on the role of SIP policies contribute new empirical evidence to a much broader literature on the ability of government interventions to reduce informational externalities. For example, a SIP mandate, which we find reduces differences in behavior across individuals with different beliefs about risk, is conceptually similar to mandates in other markets with informational externalities—for example, insurance mandates in markets with asymmetric information that aim to equalize the take-up of insurance across people with different private information about risk (Cutler et al. 2008; Einav et al. 2010; Hendren 2013; Hackmann et al. 2015).

## 2 Data and Empirical Analysis

### 2.1 Survey design

In the very early stages of the COVID-19 pandemic, we developed a survey instrument aiming to evaluate knowledge about COVID-19 and daily behaviors among people in the United States. The survey was administered by Westat, a survey research firm, between May 7 and May 26. Researchers at Westat randomly selected 13,590 residential addresses across the U.S. and mailed them an invitation to participate in an on-line survey. The addresses were selected from the Delivery Sequence File maintained by the U.S. Postal Service through an authorized vendor.

The invitation letter included a $1 cash incentive and told respondents they had the opportunity to contribute to policy development related to the COVID-19 pandemic and that they would receive $5 for completing the survey. To accommodate within-household sampling for addresses with multiple adult residents, the invitation letter randomly included instructions for the youngest male, oldest male, youngest female, or oldest female in the household to complete the survey. Alternate instructions asked either the oldest or youngest adult of the opposite sex to complete the survey if there were no males/females in the household. The invitation included a link to a website and a participant code that the respondents used to access the on-line survey. After completing the survey, the respondents were asked to provide their mailing address if they wanted to receive the $5 honorarium. Honoraria were mailed on June 5, 2020. 1,222 out of 13,590 individuals completed the survey—a response rate of 10% after accounting for non-deliverable addresses.

The survey contained questions about the risk of contracting the virus and behavioral responses to the COVID-19 pandemic across several domains, which we elicited using visual aids (Delavande 2014). Appendix A.2 shows the key questions from the survey instrument. We asked respondents to indicate whether there were directions from their governor or officials to stay or home or shelter in place. For all respondents, we asked how the time they spent on activities such as grocery shopping and dining in restaurants had changed since before the pandemic. For those responding that there were SIP restrictions active at the time of the survey, we asked how the time they spent on each activity would have changed if there had been no directions to stay at home. For those without SIP restrictions, we asked how their behaviors would have changed if restrictions had been in place. As 92% of survey respondents indicated that a SIP policy was active in their area of residence, we focus only on this group of individuals in our empirical analysis.^6^

### 2.2 Empirical Specifications

#### Heterogeneity in Subjective Risk Beliefs

Our survey instrument directly asked individuals to evaluate their risk of contracting COVID-19 (on a scale from 0% to 100%) while performing the following economic activities: 1) seeing a movie in a theater, 2) eating in a restaurant (not including take-out or delivery), 3) using shared transportation such as commercial flights, trains, buses, or shared ride services, 4) personal services such as haircuts or manicures or going to the gym, 5) grocery shopping. The activities include some that are more discretionary (seeing a movie) and some that are less discretionary (grocery shopping). We also asked survey participants about their subjective beliefs regarding the risk of outcomes conditional on infection. This set of questions included the request to assess the likelihood that individuals would (1) have symptoms; (2) need medical care; (3) need to be hospitalized; (4) die if they caught the coronavirus; (5) have (insufficient) staff and supplies for treatment in a hospital. We convert the answers to these questions into a severity index by taking an unweighted mean of the five underlying indicator variables.

We start our analysis by examining how the average level of subjective beliefs about risk compares to the level of COVID-19 prevalence reported by the Centers for Disease Control and Prevention,^7^ and the extent to which risk beliefs vary across individuals. We next ask if individuals’ subjective beliefs contain correct signals about differences in relative risk across either geographic locations or demographic groups. We use data on county-level COVID-19 prevalence, as reported by usafacts.org in May 2020, as a measure of geographic risk exposure. We use demographic and socio-economic characteristics of individuals as self-reported in the survey. The regression equations take the following form:

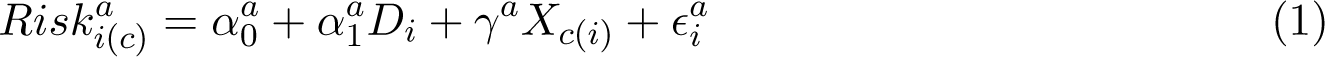

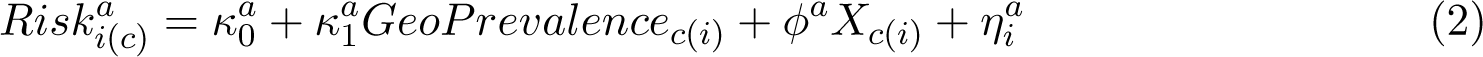

where *Risk_i(c)_^a^* is a continuous variable between 0 and 1 that measures the reported probability of contracting COVID-19 if the individual *i* residing in county *c* performed activity *a*. *D_i_* is a vector of demographic characteristics that includes: age (discretized into groups of (18-30 years, 30-44, 45-59, and 60+), sex, race (Hispanic, non-Hispanic Black, other), marital status (indicator for being married), education (indicator for having a Bachelor’s degree or above), self-reported health status (indicator for having at least one of seven chronic health conditions, including high blood pressure, diabetes, depression/anxiety, heart disease, respiratory diseases, kidney disease, and autoimmune disorder). *X_c_*_(_*_i_*_)_ is the number of days since the SIP order had been enacted. Equation 2 is an analogous specification that measures the relationship between perceived risks and the geographic prevalence of infection—*GeoPrevalence_c_*_(_*_i_*_)_ is the county-level number of COVID-19 cases per 1,000 in May 2020. The coefficients of interest are *α_1_ ^a^* and *κ_1_^a^* for each activity *a* that measure whether true variation in risk exposure across demographics or geography is predictive of individuals’ subjective risk perceptions from engaging in activity *a*. All estimating equations are weighted with survey weights and replicate jackknife weights are used to compute standard errors. See Appendix Section A.3 for details on the survey weights.

### Private Beliefs and the Choice of Preventive Behaviors

Our survey asked respondents to report how much more or less they went grocery shopping, used personal services, ate in a restaurant, saw movies in a theater, or used shared transportation since the beginning of the pandemic. The response categories for each activity included: 1) decreased a lot (by more than 50%), 2) decreased somewhat (by less than 50%), 3) has not changed, 4) increased somewhat (by less than 50%), 5) increased a lot (by more than 50%), 6) I didn’t do this before the pandemic.

For our baseline analysis of the relationship between beliefs and behaviors we create a simple indicator variable for the respondent having reduced an economic activity by a lot (more than 50%). Then, for each activity *a* we estimate a linear probability model:

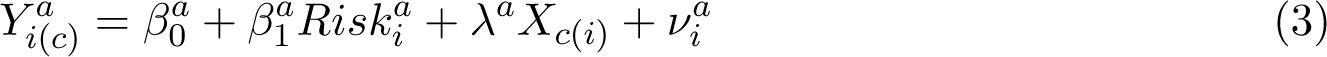

Where, *Y ^a^_i(c)_* is an indicator that takes a value of 1 of individual *i* living in county *c* reduced activity *a* by more than 50% (”by a lot”), *Risk_i_^a^* is a continuous variable between 0 and 1 that measures the subjective probability of contracting COVID-19 if the individual performed activity *a* as well as the index of beliefs about disease severity. We include the index of beliefs about disease severity in a regression for each activity *a*, as beliefs about severity conditional on contracting the disease are not activity-specific. The vector of control variables *X_c_*_(_*_i_*_)_ includes the number of days since the SIP order had been enacted to capture the idea of “SIP fatigue” that may have varied geographically depending on the timing of SIPs. Because perceptions about the underlying level of risk vary widely across individuals, *X_c_*_(_*_i_*_)_ also includes beliefs about the risk of infection when staying at home in all specifications. This adjusts for the baseline differences in risk perception levels. The coefficients of interest are *β^a^* that measure how much more or less likely individuals were to undertake an economic activity *a* if they had a one percentage point higher subjective assessment of COVID-19 risk.

We report several variations of this baseline specification in the Appendix to test the sensitivity of our results. Appendix Table A.1 reports an ordinal regression model that does not collapse responses into one indicator variable for “reducing activity by a lot.” In Appendix Table A.2 we report the results of the baseline regression using a sub-sample that excludes individuals who reported not participating in an activity before the pandemic. Appendix Table A.3 reports a specification that differences the risk of staying at home from the risk of other activities at the individual level instead of including is as the control variable. And Appendix Table A.4 reports a version of the baseline specification that includes the belief about the risk of dying only rather than the severity index. Qualitatively, the results of all these analyses are similar to the baseline.

### Perceived Role of Public Policies

To assess the role of public policies in mediating the relationship between beliefs and behaviors, we asked respondents to report how much they believe they would have changed their behavior in a hypothetical scenario without a SIP order. By comparing responses in the hypothetical scenario to the reports of the actual behavioral change, we can assess how much importance individuals ascribe to the SIP policy.

We re-estimate 150 Equation 3 using responses to the hypothetical scenario. We then compare the behavior-belief elasticity as captured by *β*_1_ between the observed and the counterfactual regime. A lower counterfactual elasticity would suggest that individuals believe that government intervention is attenuating the effect of their own risk assessment on decision-making, while a higher counterfactual elasticity would suggest that policy interventions and beliefs are complements. In the limit, a policy intervention that removes any association between subjective beliefs and behaviors (as, for instance, would be the case if all individuals were forced to stay at home) would homogenize behavioral responses of agents with heterogeneous beliefs. This intuition about the role of government mandate as an equalizer of behavioral responses across individuals with different beliefs applies to many settings outside of the infectious disease environment. For instance, a government mandate to pay into any type of social insurance program equalizes coverage purchase decisions across individuals with different beliefs about risks.

## 3 Results

### 3.1 Summary Statistics

1,365 individuals started our survey, and 1,222 individuals completed it. Our analytic sample is comprised of 1,127 individuals (92.2%) who reported being subject to an active SIP order at the time of the survey. Table 1 reports the summary statistics for this analytic sample. While the original survey was mailed to a nationally representative draw of postal addresses, which individuals decided to respond to the survey is not random.^8^ We weight the summary statistics with survey weights to account for nonrandom non-response based on observable characteristics. The average age of respondents is 47. 49% of individuals are male. 16% are Hispanic individuals while 9% are non-Hispanic Black individuals. 53% are married, 31% have a Bachelor’s degree or above, and 55% reported having at least one underlying chronic health condition among hypertension, diabetes, depression or anxiety, heart disease, a respiratory disease, kidney disease, or an autoimmune disorder. 28% reported to be an essential worker.

**Table 1:** Summary Statistics

|  | Mean | 25<sup>th</sup> pct. | 75<sup>th</sup> pct. | # of obs. |
| --- | --- | --- | --- | --- |
| Age | 47.41 | 33.00 | 62.00 | 1,089 |
| Age: <30 | 0.21 | 0.00 | 0.00 | 1,089 |
| Age: 30–44 | 0.26 | 0.00 | 1.00 | 1,089 |
| Age: 45–59 | 0.23 | 0.00 | 0.00 | 1,089 |
| Age: 60+ | 0.31 | 0.00 | 1.00 | 1,089 |
| Male | 0.49 | 0.00 | 1.00 | 1,118 |
| Non-Hispanic Black | 0.09 | 0.00 | 0.00 | 1,127 |
| Hispanic | 0.16 | 0.00 | 0.00 | 1,127 |
| Married | 0.53 | 0.00 | 1.00 | 1,120 |
| Bachelors degree+ | 0.31 | 0.00 | 1.00 | 1,121 |
| Essential worker | 0.28 | 0.00 | 1.00 | 1,117 |
| Have at least one chronic condition | 0.55 | 0.00 | 1.00 | 1,127 |
| County-level COVID-19 cases (per 1000) on May 7 | 3.38 | 0.86 | 3.10 | 1,127 |

Note: Self-reported demographic characteristics of survey participants. County-level COVID-19 cases are based May 2020 records in usafacts.org. Data source is a survey sample of the U.S. population, surveyed May 7th–26th, 2020. Statistics are weighted using survey weights that correct for non-random nonresponse. The sample is limited to 1,127 individuals, or 92.23% of survey respondents, who self-reported that a SIP order was in effect during the time of the survey.

### 3.2 Heterogeneity in Subjective Beliefs

Figure 1 plots the distributions of probability elicitations for contracting COVID-19 while engaging in different types of economic activities as well as beliefs about disease severity (Appendix Figure A.1 reports the distributions of each component of the severity index separately). Several facts are apparent from these data.

**Figure 1:**
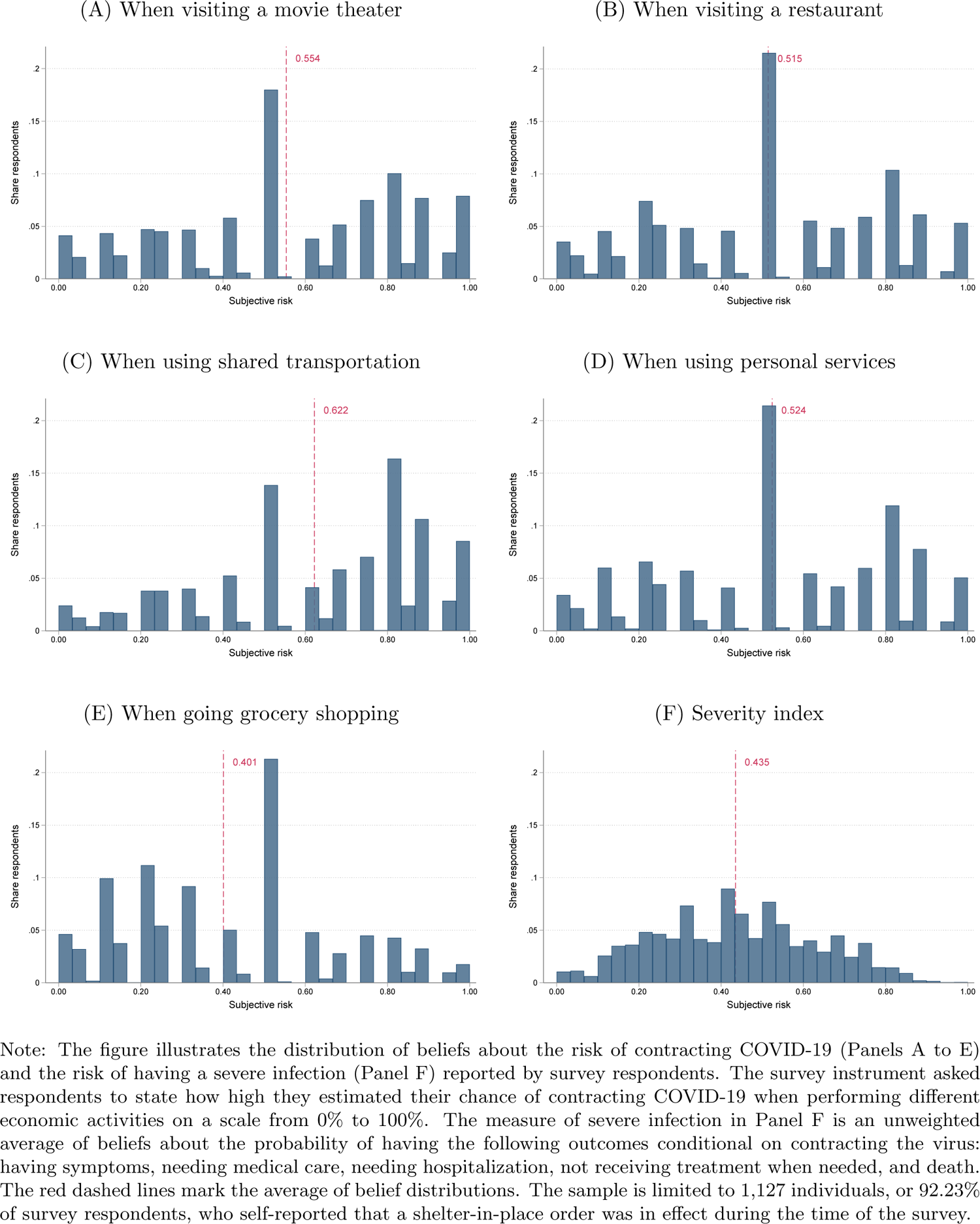
Distribution of Beliefs about Infection Risk

First, people vastly overestimated the risk of contracting COVID-19 on average (as is common in surveys eliciting subjective probabilities, see Hurd 2009; Belot et al. 2020), as well as the risk of dying from the disease.^9^ An average respondent believed there was a 40% risk of contracting COVID-19 when going grocery shopping and a 29% chance of dying from COVID conditional on contracting the virus. These belief elicitations were collected in May 2020. In May, the number of cumulative COVID-19 cases in the US was approaching 2 million (AJMC.com 2020), while the 7-day moving average of new cases oscillated between 20,000 and 25,000 people. The case fatality rate for known infections, and hence likely in itself a substantially upward biased measure of the true mortality risk, was around 6% during May 2020 (Ritchie et al. 2020). In short, the objective risk of contracting the disease was substantially lower than what survey respondents believed. Second, individual beliefs were highly heterogeneous, spanning the full support of the probability measure. As is common in the subjective belief elicitation data (Kleinjans and Soest 2014), we observe a concentration of responses around the focal point of 50% probability (15% to 30% of responses) for all categories of risk; however, substantial mass of the distribution also lies both to the left and to the right of 50%. Finally, while individuals overestimate risk levels, the within-individual *rankings* of risks were intuitive and consistent with contemporaneous public reports. As we show in Figure 4 and Appendix Figure A.2, respondents believed that staying at home was the lowest risk activity and that death was the least likely outcome.

Figure 2 examines whether heterogeneity across individuals in their risk beliefs correlates with heterogeneity in their objective risk. Appendix Table A.6 reports the tabular version of the same regression output. For each demographic variable, Panel A of Figure 2 presents the estimates of α_1_^a^ from Equation 1 for activities *a* ∈ (risk movies, restaurant, transport, service, grocery). The point estimates and 95% confidence intervals for these activities are marked in shades of blue. Several patterns emerge. First, men believe that they face a lower risk of infection than women when performing any activity. Second, Hispanic respondents believe that they face a higher risk of contracting an infection than white respondents. Non-Hispanic Black respondents also report higher perceived risk of infection (although we lack statistical power to conclude that this difference is significant). Third, people with pre-existing health conditions believe that they have a higher probability of infection. Dark grey markers and confidence intervals report the estimate of *α_1_^a^* from Equation 1 for the index measure of individuals’ beliefs about serious disease complications if infected as the outcome variable. Older individuals, Black individuals, Hispanic individuals, and those with pre-existing health conditions all believe that they are more likely to experience serious complications if infected. By contrast, individuals with a Bachelor’s degree or higher believe that they are less likely to have a more severe disease course.

**Figure 2:**
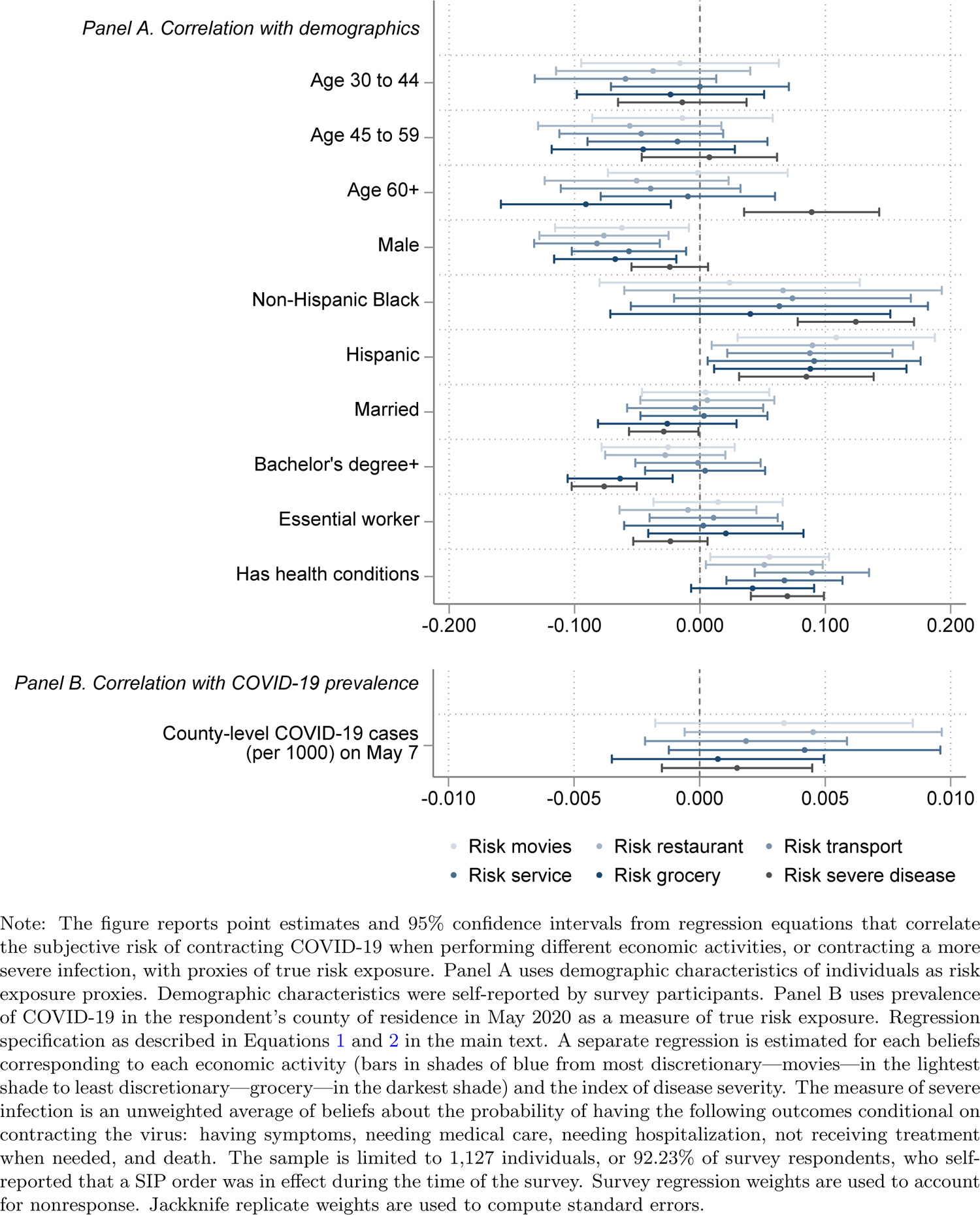
Subjective Risk Beliefs and Risk Exposure

In Panel B of Figure 2, we examine the relationship between area-level prevalence and risk beliefs. While the estimates are imprecise, they are generally positive and represent a substantive effect. For example, a change from the 25th to the 75th percentile in COVID-19 cases (2.24 per 1,000) is associated with an increase of 0.11 in the subjective probability of infection when going to a restaurant, representing a 22 percent increase relative to the mean (0.51).

In most cases, these findings are consistent with the epidemiologic evidence on the relationship between demographic characteristics and COVID-19 risk. In particular, Hispanic and Black Americans seem to be correctly aware of their heightened risk in the incidence and severity of COVID-19 relative to other demographic groups (Ogedegbe et al. 2020; Muñoz-Price et al. 2020; Van Dyke et al. 2021). The perceptions of heightened risk among racial and ethnic minorities are also consistent with other COVID-19 studies (Alsan et al. 2020; Nino et al. 2021; Reiter and Katz 2021). In the case of age, we find no evidence of a positive age gradient in the belief of infection risk, despite dramatically higher rates of recorded infection among older adults during the time period of our study (CDC 2020). Instead, those 60 and older in our sample consider themselves having a higher probability of complications conditional on infection than younger groups, consistent with observed risk (O’Driscoll et al. 2021). Our results directly contrast those of Bordalo et al. 2020 who report that older adults report a lower probability of death or hospitalization conditional on COVID-19 infection but are more consistent with those of other COVID-specific studies (Nino et al. 2021; Wise et al. 2020). Gender differences in perceived risk, in contrast, are less consistent with documented disease patterns—while men have been documented to be at a higher risk of COVID-19 infection and have worse outcomes (O’Driscoll et al. 2021), in our survey they consistently report lower risk of becoming infected than women. This type of “overconfidence” is consistent with a large body of literature documenting overconfidence in risk perceptions among men (Finucane et al. 2000) as well as studies specific to the COVID-19 pandemic (Nino et al. 2021; Dryhurst et al. 2020; Fan et al. 2020; Galasso et al. 2020). The geographic and demographic heterogeneity patterns, as well as the within-individual ranking of risks, suggest that even though individuals overestimate the overall level of risk, their subjective beliefs often contain correct signals about the direction of relative risks.

### 3.3 Private Beliefs and the Choice of Preventive Behaviors

Figure 3 presents descriptive evidence of the extent to which people reduced activities under policies restricting activity and what they reported they would have done in the absence of those policies. People reported relatively large activity reductions in the presence of a SIP order. 40 percent of people reported reducing their activity by a lot for grocery shopping, while 79 percent of people reported reducing their activity by a lot for restaurant visits, consistent with grocery shopping being a more essential activity. The figure also shows the choice of activity reductions that individuals believe they would have made absent SIP orders. The reported reduction in activity is substantially smaller than under SIP policies, but it is far from zero. Between 21% (for shared transportation) and 32% (for restaurants) of individuals report that they would have reduced the respective activities by a lot even in the absence of a formal SIP order. Thus, individuals clearly believe that SIP orders constrain their private choices, but they also believe that they would have undertaken substantial private preventive investments in the absence of government interventions.^10^

**Figure 3:**
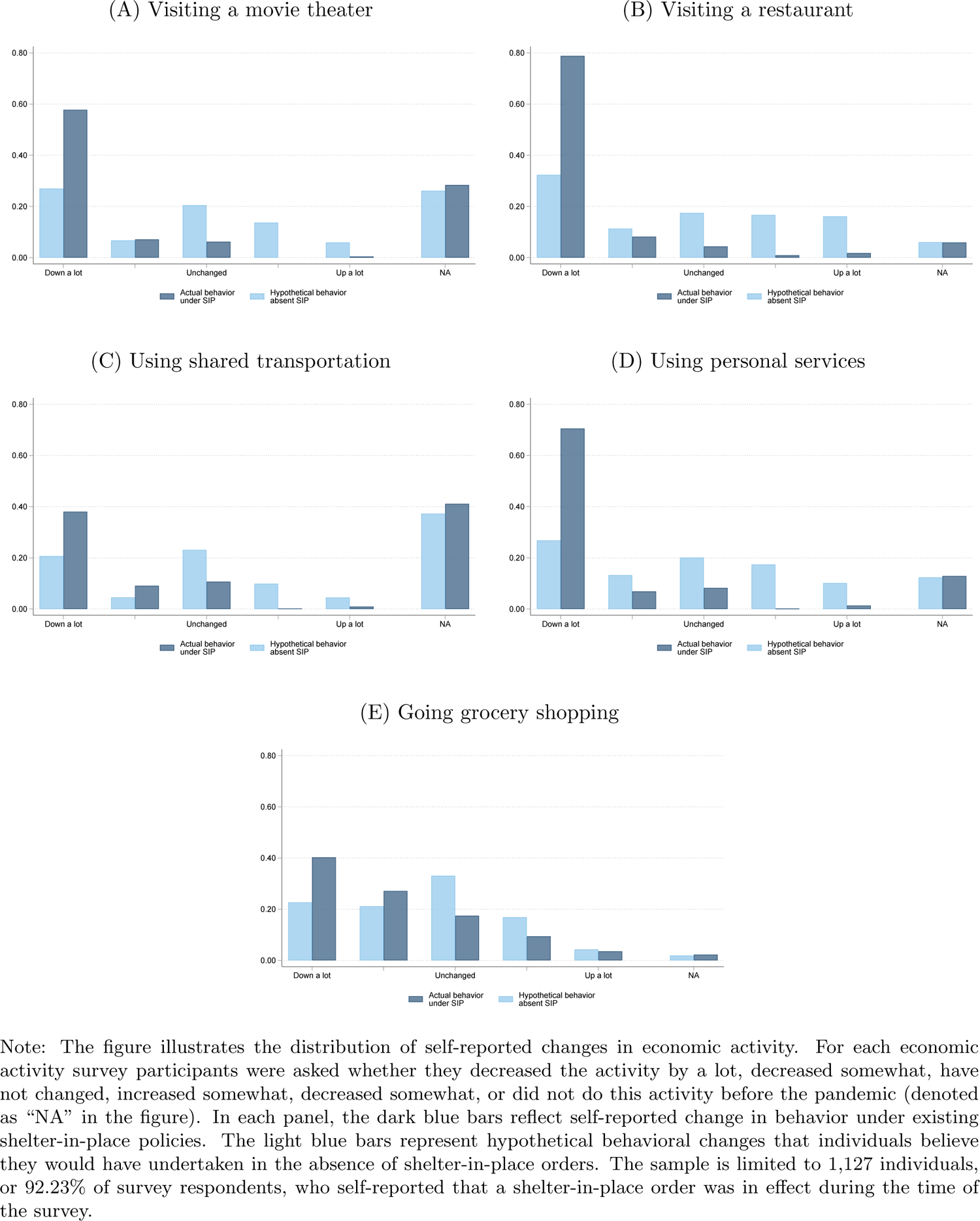
Distribution of Changes in Economic Activity

**Figure 4:**
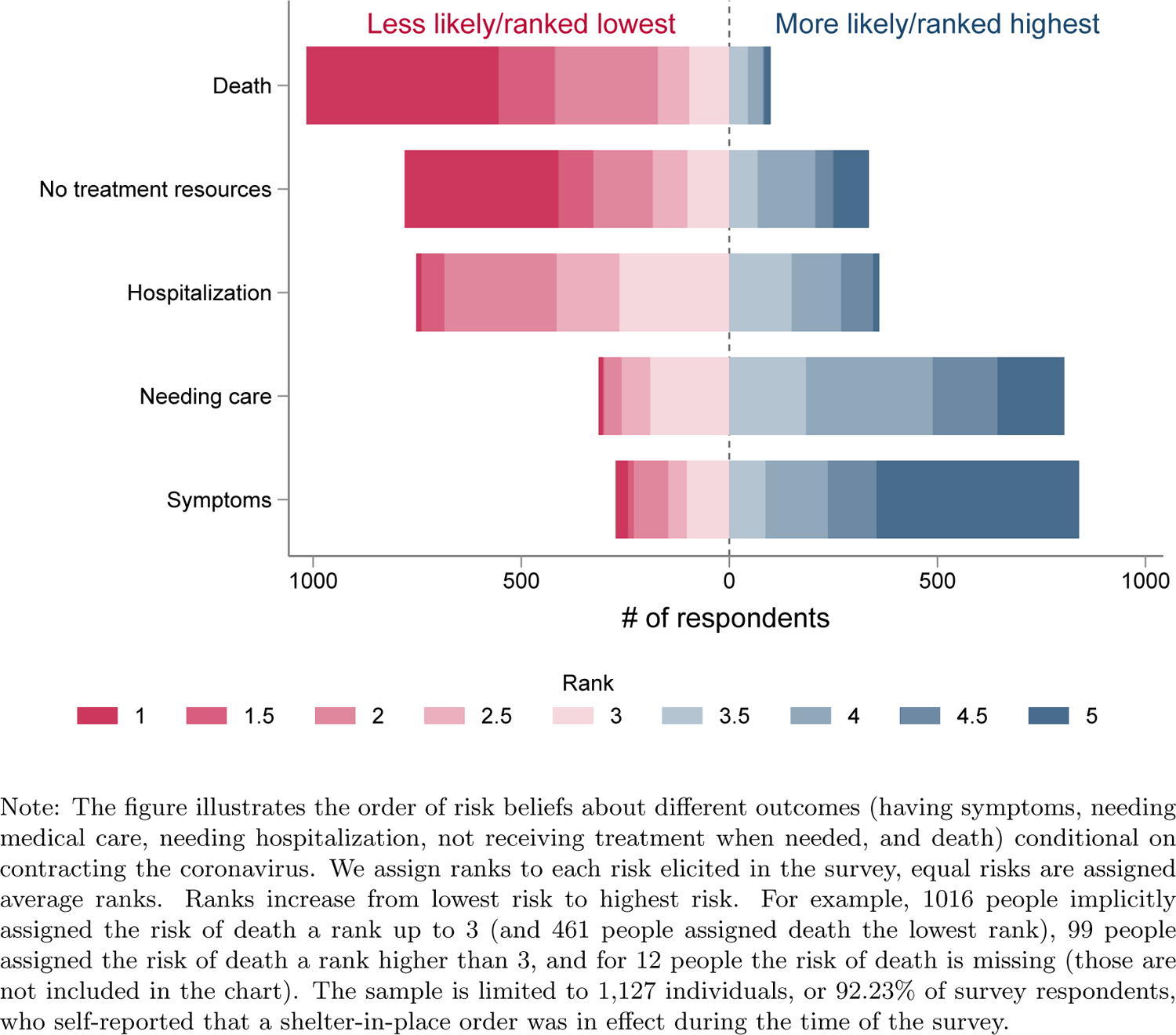
Rank Order of Beliefs about COVID-19 Severity Risk

In Table 2, we report the estimates of *β*_1_ in Equation 3 for self-reported reductions in activity under existing SIP orders. Those believing that the infection risk associated with individual activities was greater were more likely to reduce those specific activities. For example, a 10 percentage point (19% relative to the mean of 51.5 percentage points as reported in Figure 1) increase in the perceived probability of infection risk from eating in a restaurant is associated with a 2.5 percentage point increase in the probability that a respondent reduced restaurant visits by a lot (this corresponds to a 3.1% increase relative to the average probability of reducing restaurant activity by a lot, which was 79% in the sample as reported in Table 2). The estimate implies a behavior-belief elasticity of 0.16 for restaurant visits—a one percent increase in the subjective risk from a restaurant visit is associated with a 0.16 percent increase in the probability of reducing restaurant visits by a lot. The elasticity is also positive for the other activities we examined (ranging from 0.02 for movie theater to 0.20 for shared transportation), but less precise, lacking the statistical power to reject a zero elasticity at conventional confidence levels.

**Table 2:** Reduction in Economic Activity and Subjective Probability of Infection

|  | Movies |  | Restaurant |  | Transport |  | Services |  | Grocery |  |
| --- | --- | --- | --- | --- | --- | --- | --- | --- | --- | --- |
|  | Policy | No policy | Policy | No policy | Policy | No policy | Policy | No policy | Policy | No policy |
| **Subjective beliefs about risk** |  |  |  |  |  |  |  |  |  |  |
| if went to movie theater | 0.025<br>(0.072) | 0.221***<br>(0.072) |  |  |  |  |  |  |  |  |
| if went to restaurant |  |  | 0.252***<br>(0.076) | 0.425***<br>(0.095) |  |  |  |  |  |  |
| if shared transportation |  |  |  |  | 0.125<br>(0.090) | 0.149**<br>(0.073) |  |  |  |  |
| if used personal services |  |  |  |  |  |  | 0.157<br>(0.095) | 0.397***<br>(0.086) |  |  |
| if went to grocery |  |  |  |  |  |  |  |  | 0.079<br>(0.100) | 0.138<br>(0.087) |
| of more severe COVID-19 disease (index) | -0.278**<br>(0.120) | -0.026<br>(0.103) | -0.323***<br>(0.101) | 0.020<br>(0.113) | 0.009<br>(0.128) | 0.073<br>(0.105) | -0.096<br>(0.104) | 0.098<br>(0.111) | 0.444***<br>(0.114) | 0.230***<br>(0.082) |
| Mean of Dep. Var. | 0.58 | 0.27 | 0.79 | 0.32 | 0.38 | 0.21 | 0.71 | 0.27 | 0.40 | 0.23 |
| Std. Dev. of Dep. Var. | 0.49 | 0.44 | 0.41 | 0.47 | 0.49 | 0.40 | 0.45 | 0.45 | 0.49 | 0.42 |
| No. of Obs. | 1,102 | 1,099 | 1,110 | 1,108 | 1,111 | 1,104 | 1,109 | 1,111 | 1,105 | 1,109 |
| Beliefs elasticity | 0.02 | 0.45 | 0.16 | 0.68 | 0.20 | 0.44 | 0.12 | 0.77 | 0.08 | 0.24 |

Note: Table presents the results of 10 separate linear probability models as specified in Equation 3. The dependent variable is an indicator of whether the respondent decreased the economic activity by a lot. We estimate a separate model for actual self-reported change in behavior (“policy”) and self-reported change behavior in the hypothetical without SIP policies (“no policy”). The independent variables include subjective beliefs of behavior-specific risks, as well as the index of subjective severity beliefs. The measure of severe infection is an unweighted average of beliefs about the probability of having the following outcomes conditional on contracting the virus: having symptoms, needing medical care, needing hospitalization, not receiving treatment when needed, and death. Models also include the risk of staying at home and the number days since the SIP order had been enacted. The sample is limited to 1,127 individuals, or 92.23% of survey respondents, who self-reported that a SIP order was in effect during the time of the survey. Survey regression weights are used to account for nonresponse. Stanford errors using jackknife replicate weights are reported in parentheses. \* for $p < .1$ \*\* for $p < .05$ and \*\*\* for $p < .01$ .

Taken together, our results are consistent with the existence of a non-zero prevalence elasticity between the true risk and behavioral decisions (Philipson 2000; Oster 2018) that is mediated through changes in risk beliefs. Above, we found that individuals who live in areas with a higher prevalence of risk report higher subjective risk elicitations. Here, we observe that having a higher subjective risk assessment is correlated with a stronger reduction in economic activity. The two results together imply that we would expect individuals in areas with a higher prevalence of risk to undertake more preventive behaviors and reduce their economic activities more. In Appendix Table A.7 we measure the prevalence elasticity directly by correlating the geographic prevalence of COVID and the extent of economic activity reduction as reported by our survey respondents. We find that individuals in areas with a higher prevalence of COVID-19 were more likely to reduce their economic activity. The elasticity estimates (which measure the percent change in the probability of reducing an activity by a lot in response to a one percent higher prevalence of infections) range from 0.03 for movies, restaurants, and grocery shopping to 0.17 for shared transportation in the presence of government policies restricting economic activity.^11^

### 3.4 Perceived Role of Public Policies

In the second column of each column-pair in Table 2 we re-estimate the relationship between risk beliefs and privately-preferred activity reductions that respondents believe they would have chosen absent the SIP orders. In general, this counterfactual relationship is markedly stronger than the one under existing SIP orders. This is particularly true for more ‘discretionary’ services. For example, the point estimate for the relationship between risk beliefs and behavior is more than twice as large for the use of personal services in the absence of SIP (0.40) than when SIP is in place (0.16). Differences in the estimates of the relationship between perceived risk of infection and behavior reduction are statistically significant with a p-value for a two-sided test of *<* 0.1 for movies, restaurants, and services.^12^

As with belief-behavior relationship, the relationship between geographic prevalence of disease and activity reduction is stronger for the hypothetical scenario of no policy intervention. We estimate higher hypothetical prevalence elasticities for all economic activities, ranging from 0.11 for restaurants and personal services to 0.21 for shared transportation. Higher belief-behavior and prevalence elasticities that we estimate for the counterfactual scenarios without “directions from your governor or other officials to stay at home or shelter-in-place” orders are consistent with individuals believing that public policy interventions are in fact constraining their private choices. Respondents believe that in the absence of SIP policies their own subjective beliefs about risk would have played a more important role in driving their behavioral choices.

### 3.5 Risk of Infection versus Risk of Severe Disease

The relationship between the perceived risk of a poor outcome conditional on infection and activity reductions also differs depending on whether SIP is in place. In the absence of SIP policies, we find no statistically significant association between activity reductions and beliefs regarding risk of complications for each activity, with the exception of grocery shopping. For grocery shopping, people who believe they are likely to experience complications if infected are more likely than those who perceive the risk to be low to reduce this activity a lot. By contrast, in the presence of SIP policies, those who believe that they face a greater risk of a serious complication if infected are less likely to reduce several activities by a lot. In other words, those at high risk of a poor outcome conditional on infection are more likely to maintain or increase the extent to which they go to a sit down restaurant and go to a movie theater than those who perceive themselves at lower risk. This is consistent with people with high perceived health risk reducing their protective behaviors relative to those with lower perceived risk when policies create a less risky environment. While this relationship differs for grocery shopping, we note that grocery shopping is typically exempt from SIP policies. Thus, the implementation of SIP is unlikely to reduce the risk associated with this activity.

## 4 Conclusion

We find that subjective assessments of risk are strongly predictive of economic activity. Our survey results suggest that a large share of Americans dramatically reduced their activities outside the home in response to the COVID-19 pandemic, and that perceptions of risk influenced these choices. Individuals varied in their perceptions of both the risk of infection and the risk of a poor outcome conditional on infection early in the COVID-19 pandemic. Given the crucial role of private beliefs in shaping behavior, an important question is how individual beliefs are formed and whether they reflect reality. Unlike prior work in the context of COVID-19 (Bordalo et al. 2020), we find that differences in the perception of risks across demographic subgroups and different geographies generally correlate with known realities of the risk of contracting the disease (Benitez et al. 2020; Ford et al. 2020; Richardson et al. 2020; Polyakova et al. 2020; Polyakova et al. 2021). This finding of a positive relationship between risk perceptions and protective behaviors in the context of the COVID-19 pandemic is consistent with other contemporaneous studies (Dryhurst et al. 2020; Heffetz and Ishai 2021).

An implication of our results is that SIP policies equalize behavior restrictions, to some extent, across those with more disparate underlying beliefs about disease risk. In particular, our results imply that formal SIP orders are likely to be more constraining (relative to the level of activities that individuals would have chosen privately) for people who believe they have (and often indeed have) a lower risk of contracting a COVID-19 infection. We found that many respondents believe that they would have dramatically reduced their activities even in the absence of formal policies restricting such activities. These findings imply that epidemiologic models of SIP effects are likely to overestimate the effectiveness of policy interventions, as individuals are likely to alter their behaviors even in the absence of interventions based on their beliefs about risks. More generally our results indicate that when designing policies that aim to change individual behavior in the presence of risk, policy makers need to consider how subjective risk perceptions are formed and how they shape behavior.

## Data Availability

All data produced in the present study are available upon reasonable request to the authors

## A Appendix

### A.1 Literature on subjective risk beliefs measurement

Research in both low- and high-income countries has examined aspects of subjective probability measurement, frequently implemented in surveys. Several generalizations emerge from this research that relate to the elicitations of subjective probabilities reported in this paper:

- Studies in high-income countries, and more recently in lower-income countries, generally conclude that survey respondents (even those with less education) can understand and answer probabilistic questions (Manski 2004; Delavande 2014).
- Elicited probabilistic expectations have also been generally found to follow basic properties of probabilities (Delavande et al. 2011).
- The specific method used to measure subjective probabilities matters (Attanasio 2009), but in high-income countries, the standard approach (used in the HRS, for example) is to ask about perceived probabilities in a percent chance format (Manski 2004; Hurd 2009), which is similar to our approach. Visual aids accompanying such questions (as we use) have also been demonstrated to be valuable (Delavande 2014).
- Subjective probability measures can be biased and can suffer from the use of focal-point heuristics:
  **–** Subjective probabilities measured through household surveys generally contain a high frequency of focal point responses (0, 50, and 100, for example) (Fischhoff and Bruine De Bruin 1999; Bruin et al. 2000; Hurd 2009). This is true across many different substantive topics of research (Hurd 2009).
  **–** Pooling across different topics studied using subjective probabilities (mortality/survival, retirement, and stock market performance), when the actual probability of an event is greater than 50%, subjective probabilities are generally understated, while when the actual probability of an event is less than 50% subjective probabilities are generally overstated (Hurd 2009)—as we also generally observe in our data.
  **–** Respondents often overestimate the probability of events that are more salient (Viscusi et al. 1998).
  Nonetheless, and importantly, subjective probability measures have been shown to predict actual future decisions and behavior (Finkelstein and McGarry 2006; Dominitz and Manski 2007; Hurd 2009; Delavande et al. 2011).
  Subjective probabilities have also been shown to vary with observable characteristics in the same way that actual outcomes do Hurd and McGarry 1995—but heterogeneity in beliefs across individuals generally exists too (beliefs which should not necessarily require private, individual-specific information) (Manski 2004; Hurd 2009; Delavande 2014).
- On subjective probability measurement specific to health/mortality:

**–** Mortality expectations have been shown to vary with observable characteristics and risk factors in expected ways—for example, with age, time horizon, education, and even HIV status not known to the respondents at the time of the survey (Hamermesh 1985; Delavande and Kohler 2009).
**–** Subjective survival probabilities also correlate with life table data (Hurd and McGarry 1995).
**–** And subjective survival probabilities have even been shown to be predictive of actual survival (Hurd and McGarry 2002).
**–** In the case of COVID-19, research has found that individuals tend to overestimate the health risk (present paper, as well as Akesson et al. (2020) and Heffetz and Ishai (2021))

### A.2 Survey questionnaire

#### (1) Questions about Behavioral Changes

How has the time you spend on each of the following activities changed since the pandemic?

##### Q3A Going to the grocery store

1. Decreased a lot (by more than 50%)
2. Decreased somewhat (by less than 50%)
3. Has not changed
4. Increased somewhat (by less than 50%)
5. Increased a lot (by more than 50%)
6. I didn’t do this before the pandemic

##### Q3B Receive personal services such as haircuts or manicures, or go to the gym

1. Decreased a lot (by more than 50%)
2. Decreased somewhat (by less than 50%)
3. Has not changed
4. Increased somewhat (by less than 50%)
5. Increased a lot (by more than 50%)
6. I didn’t do this before the pandemic

##### Q3C Exercise outdoors

1. Decreased a lot (by more than 50%)
2. Decreased somewhat (by less than 50%)
3. Has not changed
4. Increased somewhat (by less than 50%)
5. Increased a lot (by more than 50%)
6. I didn’t do this before the pandemic

##### Q3D Eat in a restaurant (not including take out or delivery)

1. Decreased a lot (by more than 50%)
2. Decreased somewhat (by less than 50%)
3. Has not changed
4. Increased somewhat (by less than 50%)
5. Increased a lot (by more than 50%)
6. I didn’t do this before the pandemic

##### Q3F See a movie in a theater

1. Decreased a lot (by more than 50%)
2. Decreased somewhat (by less than 50%)
3. Has not changed
4. Increased somewhat (by less than 50%)
5. Increased a lot (by more than 50%)
6. I didn’t do this before the pandemic

##### Q3G Use shared transportation (such as commercial flights, trains, buses, or shared ride services)

1. Decreased a lot (by more than 50%)
2. Decreased somewhat (by less than 50%)
3. Has not changed
4. Increased somewhat (by less than 50%)
5. Increased a lot (by more than 50%)
6. I didn’t do this before the pandemic

#### (2) Elicitation of Subjective Beliefs about Risk

##### The next questions ask about the chances of you or someone like you catching the coronavirus

Q10 How likely is it that you or someone like you would catch the coronavirus if you stayed at home the vast majority of the time? (Percentage 0-100)

0 (No chance) – 100 (Sure to happen)

Q11 How likely is it that you or someone like you would catch the coronavirus if you received personal services, such as haircuts or manicures, or went to the gym?

0 (No chance) – 100 (Sure to happen)

Q12 How likely is it that you or someone like you would catch the coronavirus if you exercised outdoors?

0 (No chance) – 100 (Sure to happen)

Q13 How likely is it that you or someone like you would catch the coronavirus if you went to the grocery store?

0 (No chance) – 100 (Sure to happen)

Q14 How likely is it that you or someone like you would catch the coronavirus if you ate at a restaurant, not including take out or delivery?

0 (No chance) – 100 (Sure to happen)

Q15 How likely is it that you or someone like you would catch the coronavirus if you went to work regularly outside your home?

0 (No chance) – 100 (Sure to happen)

Q16 How likely is it that you or someone like you would catch the coronavirus if you saw a movie in a theater?

0 (No chance) – 100 (Sure to happen)

Q17 How likely is it that you or someone like you would catch the coronavirus if you used shared transportation, such as commercial flights, trains, buses, or shared ride services?

0 (No chance) – 100 (Sure to happen)

##### The next questions ask about what would happen if you or somebody like you caught the coronavirus

Q18 If you or somebody like you caught the coronavirus, how likely is it that you would not have any symptoms?

0 (No chance) – 100 (Sure to happen)

Q19 If you or somebody like you caught the coronavirus, how likely is it that you would need medical care?

0 (No chance) – 100 (Sure to happen)

Q20 If you or somebody like you caught the coronavirus, how likely is it that you would need to be hospitalized?

0 (No chance) – 100 (Sure to happen)

Q21 If you or somebody like you caught the coronavirus, how likely is it that you would die?

0 (No chance) – 100 (Sure to happen)

Q22 If you or somebody like you caught the coronavirus, how likely is it that the hospital would have the staff and supplies to treat you?

0 (No chance) – 100 (Sure to happen)

### A.3 Survey weights

We use survey weights to account for non-random nonresponse based on observable demographics in all analyses.

The computation of the survey weights began with the base weights, which were then adjusted for differential nonresponse to the survey request and to within-household selection, and then calibrated to external totals. The base weight is the reciprocal of the probability of selection of the address. The adjustment for nonresponse used a weighting class adjustment to redistribute base weights of eligible nonrespondents to eligible respondents within the same weighting class, after accounting for the estimated proportion of cases with undetermined eligibility that are eligible; the weighting classes were defined using address-level variables available on the sampling frame and census tract-level characteristics from the American Community Survey (ACS; 2018 5-year tables). Candidate variables included an indicator of whether a name could be matched to the address, the dwelling type (multi-unit structure or not), census region, indicators (from USPS files) of whether the address is vacant and whether the address is seasonal, and quartiles of the following census tract-level characteristics: percent below poverty, percent with less than a high school diploma, percent with a college degree or higher, percent age 65+, percent Black, and percent Hispanic. A classification tree algorithm was used to identify the classes, with survey response status as the variable being modeled.

Next, the nonresponse adjusted weights were adjusted to account for the selection of one adult among the adults in the household. The adjustment factor is the number of adults in the household. Finally, the adjusted weights were raked to population estimates from the ACS (2018; 1-year tables). This raking adjustment aligns estimated totals from the survey to the ACS estimates on the following dimensions: (1) Sex by age category (18-29, 30-49, 50-69, 70+); (2) Race (White alone, Black alone, other) of persons age 18 or older, by census region; and (3) Ethnicity (Hispanic, non-Hispanic) of persons age 18 or older, by census region.

For computing basic descriptive statistic point estimates, the survey weights themselves are sufficient to account for the complex sample design. But for estimating the precision of those estimates (e.g., producing standard errors and confidence intervals), it is necessary to use a method that takes into account the precision effects of the complex sampling and estimation procedures used in this study. The method we used was to compute replicate weights using the unstratified “delete one group” jackknife with 80 random groups. We constructed the replicates by randomly sorting the sampled addresses into 80 groups and then deleting one group at a time, to result in 80 replicates. For each replicate, a set of replicate base weights is produced by first assigning a replicate weight multiplier of 0 to the addresses that were deleted in constructing the replicate and assigning a replicate weight multiplier of 80/79 to the addresses that were not deleted in constructing the replicate, then multiplying the full-sample address base weight by the replicate weight multiplier. Each set of replicate weights underwent the same set of adjustments that were applied to the full-sample weights.

## A.4 Appendix Figures and Tables

**Figure A.1:**
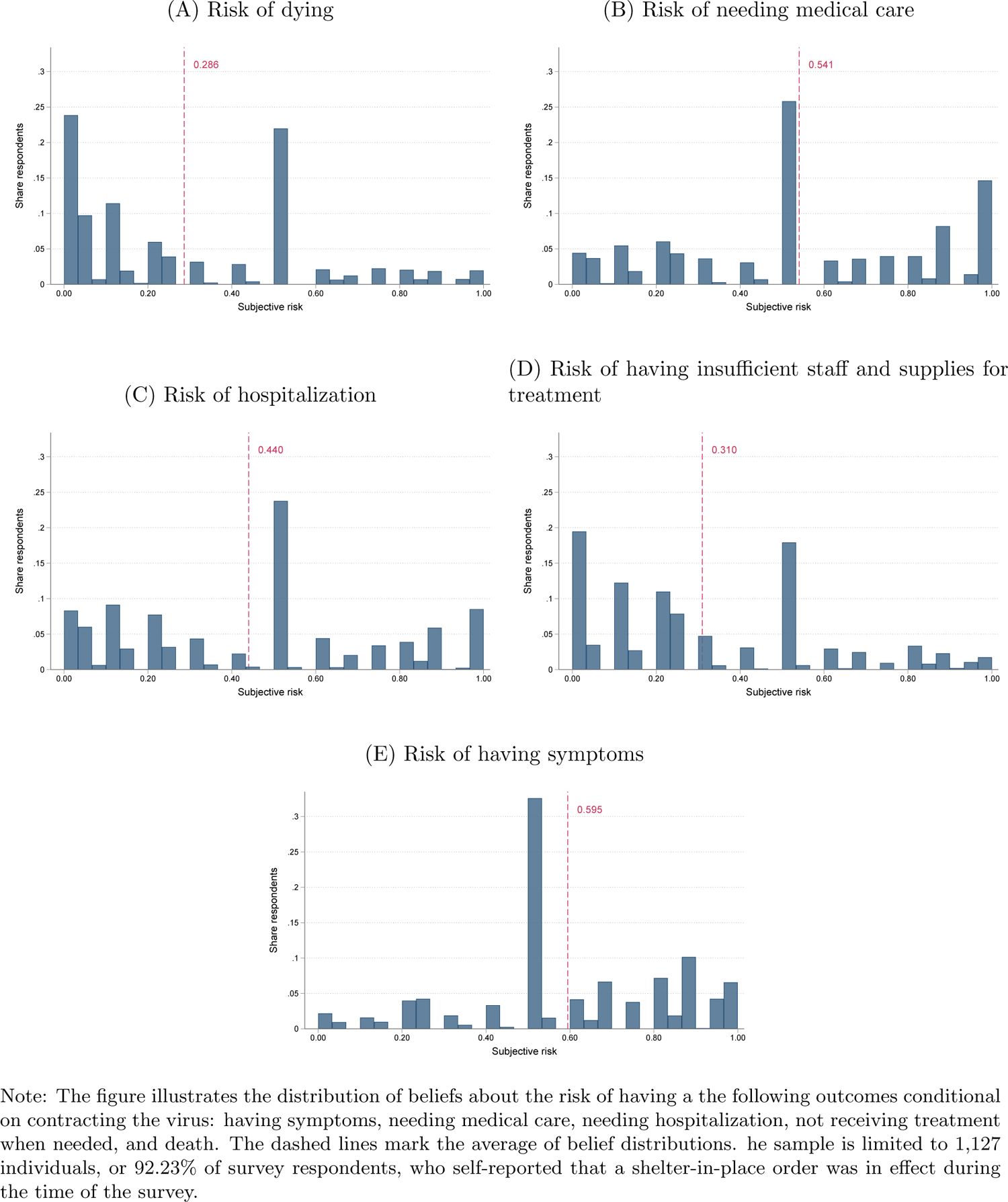
Distribution of Beliefs about COVID-19 Severity Risk

**Figure A.2:**
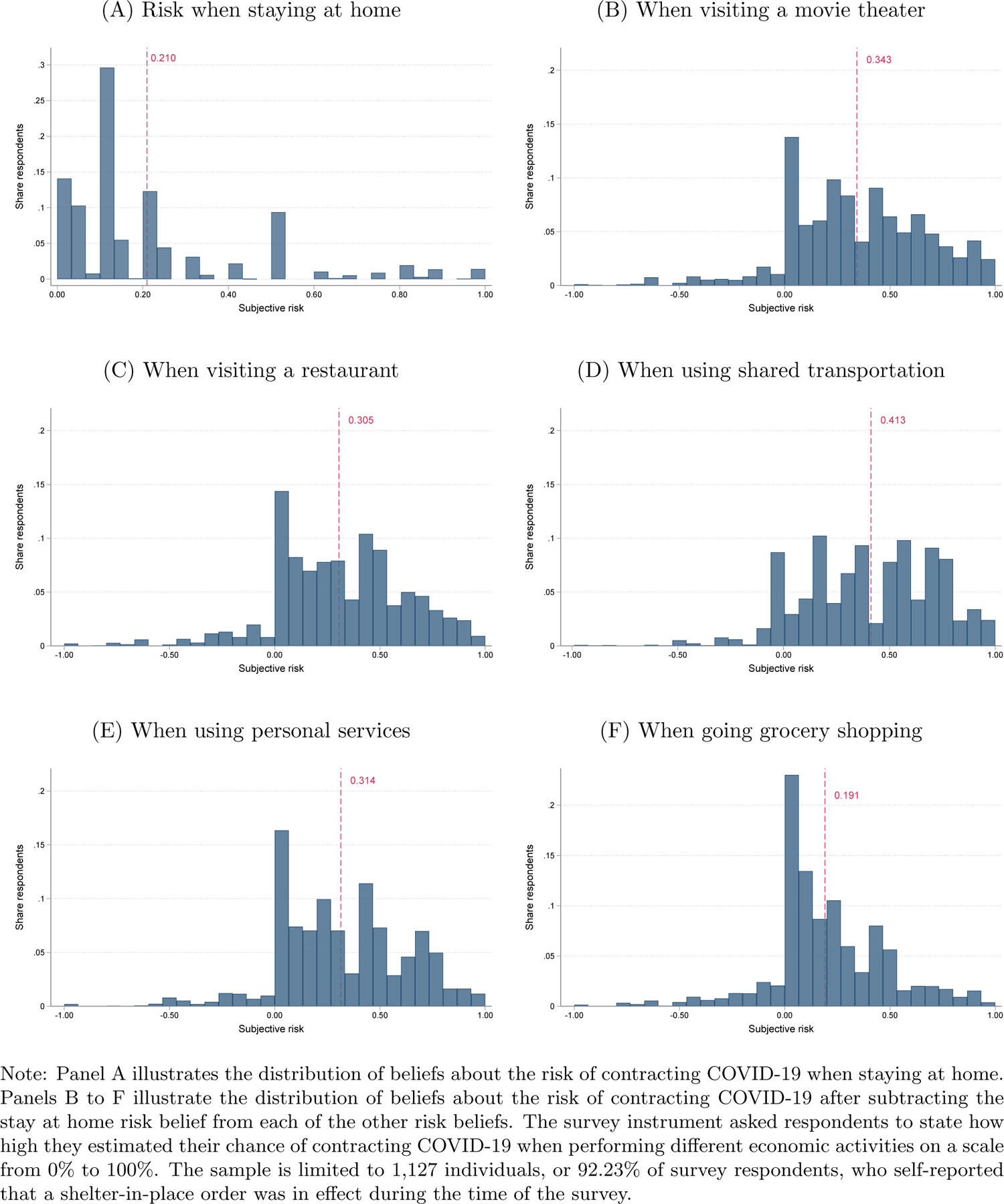
Distribution of Beliefs about Infection Risk after Subtracting Beliefs about Risk when Staying Home

**Table A.1:** Reduction in Economic Activity and Subjective Probability of Infection - Ordered Logit

|  | Movies |  | Restaurant |  | Transport |  | Services |  | Grocery |  |
| --- | --- | --- | --- | --- | --- | --- | --- | --- | --- | --- |
|  | Policy | No policy | Policy | No policy | Policy | No policy | Policy | No policy | Policy | No policy |
| **Subjective beliefs about risk** |  |  |  |  |  |  |  |  |  |  |
| *if went to movie theater* |  |  |  |  |  |  |  |  |  |  |
| predict "increased a lot" | -0.000<br>(0.001) | -0.055***<br>(0.019) |  |  |  |  |  |  |  |  |
| predict "increased somewhat" |  | -0.104***<br>(0.037) |  |  |  |  |  |  |  |  |
| predict "unchanged" | -0.020<br>(0.065) | -0.065**<br>(0.026) |  |  |  |  |  |  |  |  |
| predict "decreased somewhat" | - 0.002<br>(0.005) | 0.027**<br>(0.011) |  |  |  |  |  |  |  |  |
| predict "decreased a lot" | 0.022<br>(0.072) | 0.197***<br>(0.064) |  |  |  |  |  |  |  |  |
| *if went to restaurant* |  |  |  |  |  |  |  |  |  |  |
| predict "increased a lot" |  |  | -0.025**<br>(0.012) | -0.143***<br>(0.046) |  |  |  |  |  |  |
| predict "increased somewhat" |  |  | -0.010*<br>(0.006) | -0.097**<br>(0.037) |  |  |  |  |  |  |
| predict "unchanged" |  |  | -0.136***<br>(0.041) | -0.032*<br>(0.018) |  |  |  |  |  |  |
| predict "decreased somewhat" |  |  | -0.090***<br>(0.032) | 0.030**<br>(0.012) |  |  |  |  |  |  |
| predict "decreased a lot" |  |  | 0.262***<br>(0.078) | 0.241***<br>(0.085) |  |  |  |  |  |  |

| *(Cont'd.)* | Movies |  | Restaurant |  | Transport |  | Services |  | Grocery |  |
| --- | --- | --- | --- | --- | --- | --- | --- | --- | --- | --- |
|  | Policy | No policy | Policy | No policy | Policy | No policy | Policy | No policy | Policy | No policy |
| ***if shared transportation*** |  |  |  |  |  |  |  |  |  |  |
| predict "increased a lot" |  |  |  |  | -0.003<br>(0.004) | -0.028**<br>(0.013) |  |  |  |  |
| predict "increased somewhat" |  |  |  |  | -0.001<br>(0.001) | -0.055*<br>(0.028) |  |  |  |  |
| predict "unchanged" |  |  |  |  | -0.082<br>(0.087) | -0.044<br>(0.027) |  |  |  |  |
| predict "decreased somewhat" |  |  |  |  | 0.004<br>(0.005) | 0.017*<br>(0.009) |  |  |  |  |
| predict "decreased a lot" |  |  |  |  | 0.081<br>(0.086) | 0.110*<br>(0.056) |  |  |  |  |
| ***if used personal services*** |  |  |  |  |  |  |  |  |  |  |
| predict "increased a lot" |  |  |  |  |  |  | -0.009<br>(0.007) | -0.118***<br>(0.028) |  |  |
| predict "increased somewhat" |  |  |  |  |  |  | -0.001<br>(0.001) | -0.154***<br>(0.041) |  |  |
| predict "unchanged" |  |  |  |  |  |  | -0.109<br>(0.076) | -0.068**<br>(0.028) |  |  |
| predict "decreased somewhat" |  |  |  |  |  |  | -0.024<br>(0.018) | 0.064***<br>(0.018) |  |  |
| predict "decreased a lot" |  |  |  |  |  |  | 0.144<br>(0.100) | 0.276***<br>(0.073) |  |  |

*(continued on next page)*

| *(Cont'd.)* | Movies |  | Restaurant |  | Transport |  | Services |  | Grocery |  |
| --- | --- | --- | --- | --- | --- | --- | --- | --- | --- | --- |
|  | Policy | No policy | Policy | No policy | Policy | No policy | Policy | No policy | Policy | No policy |
| ***if went to grocery*** |  |  |  |  |  |  |  |  |  |  |
| predict "increased a lot" |  |  |  |  |  |  |  |  | -0.012<br>(0.013) | -0.010<br>(0.013) |
| predict "increased somewhat" |  |  |  |  |  |  |  |  | -0.028<br>(0.029) | -0.032<br>(0.045) |
| predict "unchanged" |  |  |  |  |  |  |  |  | -0.041<br>(0.046) | -0.021<br>(0.030) |
| predict "decreased somewhat" |  |  |  |  |  |  |  |  | -0.009<br>(0.011) | 0.018<br>(0.025) |
| predict "decreased a lot" |  |  |  |  |  |  |  |  | 0.090<br>(0.098) | 0.045<br>(0.064) |
| ***of more severe COVID-19 disease (index)*** |  |  |  |  |  |  |  |  |  |  |
| predict "increased a lot" | 0.006<br>(0.007) | -0.014<br>(0.025) | 0.033**<br>(0.015) | -0.069<br>(0.059) | 0.002<br>(0.005) | -0.001<br>(0.021) | 0.009<br>(0.008) | -0.086**<br>(0.037) | -0.043***<br>(0.015) | -0.034**<br>(0.017) |
| predict "increased somewhat" |  | -0.026<br>(0.048) | 0.013<br>(0.008) | -0.047<br>(0.041) | 0.000<br>(0.001) | -0.001<br>(0.040) | 0.001<br>(0.001) | -0.113**<br>(0.047) | -0.097***<br>(0.032) | -0.109**<br>(0.051) |
| predict "unchanged" | 0.290**<br>(0.111) | -0.016<br>(0.030) | 0.175***<br>(0.052) | -0.015<br>(0.014) | 0.056<br>(0.122) | -0.001<br>(0.033) | 0.106<br>(0.078) | -0.050**<br>(0.021) | -0.143***<br>(0.048) | -0.071**<br>(0.034) |
| predict "decreased somewhat" | 0.022**<br>(0.009) | 0.007<br>(0.012) | 0.116***<br>(0.038) | 0.015<br>(0.013) | -0.003<br>(0.007) | 0.000<br>(0.013) | 0.024<br>(0.017) | 0.047**<br>(0.021) | -0.031*<br>(0.019) | 0.061**<br>(0.029) |
| predict "decreased a lot" | -0.318**<br>(0.121) | 0.049<br>(0.090) | -0.336***<br>(0.098) | 0.116<br>(0.100) | -0.055<br>(0.121) | 0.002<br>(0.081) | -0.139<br>(0.101) | 0.202**<br>(0.081) | 0.315***<br>(0.102) | 0.153**<br>(0.071) |
| Mean of Dep. Var. | 1.78 | 2.66 | 1.39 | 2.73 | 2.16 | 2.73 | 1.54 | 2.70 | 2.09 | 2.58 |
| Std. Dev. of Dep. Var. | 0.96 | 1.19 | 0.84 | 1.46 | 0.98 | 1.04 | 0.91 | 1.30 | 1.13 | 1.14 |
| No. of Obs. | 1,102 | 1,099 | 1,110 | 1,108 | 1,111 | 1,104 | 1,109 | 1,111 | 1,105 | 1,109 |

Note: Table reports the result of re-estimating the models in Table 2 of the main manuscript using an ordered logit instead of a linear probability model.

**Table A.2:** Reduction in Economic Activity and Subjective Probability of Infection - Drop Individuals Reporting They Did Not Participate in an Activity Before the Pandemic

|  | Movies |  | Restaurant |  | Transport |  | Services |  | Grocery |  |
| --- | --- | --- | --- | --- | --- | --- | --- | --- | --- | --- |
|  | Policy | No policy | Policy | No policy | Policy | No policy | Policy | No policy | Policy | No policy |
| **Subjective beliefs about risk** |  |  |  |  |  |  |  |  |  |  |
| if went to movie theater | 0.077<br>(0.081) | 0.339***<br>(0.096) |  |  |  |  |  |  |  |  |
| if went to restaurant |  |  | 0.264***<br>(0.075) | 0.437***<br>(0.101) |  |  |  |  |  |  |
| if shared transportation |  |  |  |  | 0.330***<br>(0.112) | 0.295**<br>(0.112) |  |  |  |  |
| if used personal services |  |  |  |  |  |  | 0.182**<br>(0.085) | 0.470***<br>(0.093) |  |  |
| if went to grocery |  |  |  |  |  |  |  |  | 0.082<br>(0.102) | 0.142<br>(0.089) |
| of more severe COVID-19 disease (index) | -0.124<br>(0.124) | 0.073<br>(0.143) | -0.251**<br>(0.097) | 0.058<br>(0.119) | 0.094<br>(0.151) | 0.073<br>(0.151) | -0.009<br>(0.106) | 0.104<br>(0.126) | 0.477***<br>(0.111) | 0.247***<br>(0.084) |
| Mean of Dep. Var. | 0.81 | 0.37 | 0.84 | 0.34 | 0.65 | 0.32 | 0.82 | 0.30 | 0.41 | 0.23 |
| Std. Dev. of Dep. Var. | 0.39 | 0.48 | 0.37 | 0.47 | 0.48 | 0.47 | 0.39 | 0.46 | 0.49 | 0.42 |
| No. of Obs. | 796 | 795 | 1,057 | 1,055 | 725 | 719 | 986 | 989 | 1,089 | 1,094 |

Note: Table reports the result of re-estimating the models in Table 2 excluding observations for individuals who responded “I did not do this before the pandemic” from the estimation sample.

**Table A.3:**
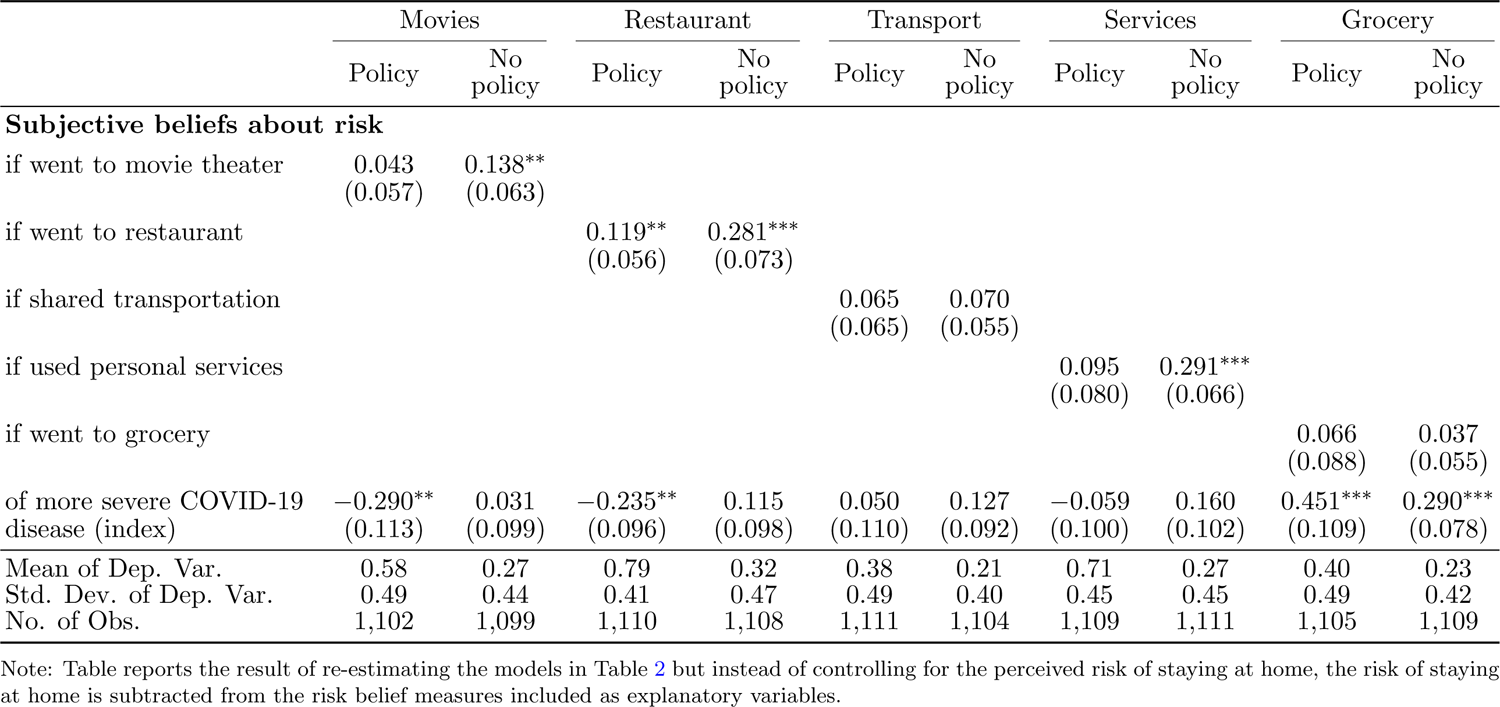
Reduction in Economic Activity and Subjective Probability of Infection - Subtracting Beliefs about the Risk when Staying at Home from Beliefs about Risks from other Activities

**Table A.4:** Reduction in Economic Activity and Subjective Probability of Infection - Severity Index Replaces with the Risk of Dying Only

|  | Movies |  | Restaurant |  | Transport |  | Services |  | Grocery |  |
| --- | --- | --- | --- | --- | --- | --- | --- | --- | --- | --- |
|  | Policy | No policy | Policy | No policy | Policy | No policy | Policy | No policy | Policy | No policy |
| **Subjective beliefs about risk** |  |  |  |  |  |  |  |  |  |  |
| if went to movie theater | 0.031<br>(0.072) | 0.240***<br>(0.069) |  |  |  |  |  |  |  |  |
| if went to restaurant |  |  | 0.274***<br>(0.079) | 0.459***<br>(0.088) |  |  |  |  |  |  |
| if shared transportation |  |  |  |  | 0.166*<br>(0.088) | 0.183**<br>(0.071) |  |  |  |  |
| if used personal services |  |  |  |  |  |  | 0.143<br>(0.094) | 0.424***<br>(0.082) |  |  |
| if went to grocery |  |  |  |  |  |  |  |  | 0.105<br>(0.105) | 0.144<br>(0.089) |
| of dying from COVID-19 | -0.233**<br>(0.089) | -0.080<br>(0.069) | -0.315***<br>(0.078) | -0.046<br>(0.078) | -0.107<br>(0.085) | -0.018<br>(0.071) | -0.076<br>(0.076) | 0.011<br>(0.078) | 0.271***<br>(0.088) | 0.158**<br>(0.077) |
| Mean of Dep. Var. | 0.58 | 0.27 | 0.79 | 0.32 | 0.39 | 0.20 | 0.71 | 0.27 | 0.40 | 0.23 |
| Std. Dev. of Dep. Var. | 0.49 | 0.44 | 0.41 | 0.47 | 0.49 | 0.40 | 0.45 | 0.44 | 0.49 | 0.42 |
| No. of Obs. | 1,094 | 1,091 | 1,101 | 1,101 | 1,103 | 1,097 | 1,100 | 1,103 | 1,096 | 1,101 |

Note: Table reports the result of re-estimating the models in Table 2 but instead of the COVID-19 severity index, it includes only includes the perceived risk of dying from COVID-19 conditional on catching COVID-19.

**Table A.5:** Selection into Survey Completion

| | Completed | | Not completed | | $\Delta$ | $t\text{-stat}$ | $p$ |
| --- | --- | --- | --- | --- | --- | --- | --- |
|  | Mean | SD | Mean | SD |  |  |  |
| *Race/ethnicity* |  |  |  |  |  |  |  |
| Share Non-Hispanic Black | 0.08 | 0.14 | 0.13 | 0.20 | −0.05 | 11.08 | 0.00 |
| Share Hispanic | 0.13 | 0.17 | 0.15 | 0.20 | −0.02 | 4.94 | 0.00 |
| *Education* |  |  |  |  |  |  |  |
| Share with less than GED | 0.05 | 0.04 | 0.07 | 0.05 | −0.02 | 11.54 | 0.00 |
| Share with GED or equivalent | 0.22 | 0.13 | 0.25 | 0.13 | −0.03 | 6.84 | 0.00 |
| Share with Bachelors degree+ | 0.28 | 0.19 | 0.24 | 0.17 | 0.04 | −8.11 | 0.00 |
| *Income* |  |  |  |  |  |  |  |
| Median income (log) | 9.95 | 1.66 | 9.87 | 1.62 | 0.08 | −1.60 | 0.11 |
| Share below poverty line | 0.11 | 0.09 | 0.13 | 0.11 | −0.02 | 9.15 | 0.00 |
| Share in labor force | 0.59 | 0.20 | 0.58 | 0.19 | 0.01 | −1.45 | 0.15 |
| *Other demographics* |  |  |  |  |  |  |  |
| Share of non-citizens | 0.06 | 0.07 | 0.06 | 0.08 | 0.00 | 3.58 | 0.00 |
| Share with children under 18 | 0.38 | 0.16 | 0.38 | 0.15 | 0.00 | 1.23 | 0.22 |
| Share over age 65 | 0.15 | 0.09 | 0.14 | 0.09 | 0.01 | −1.33 | 0.18 |
| Share with a disability | 0.09 | 0.05 | 0.10 | 0.06 | −0.01 | 6.44 | 0.00 |
| Share of buildings occupied by owners | 0.63 | 0.27 | 0.58 | 0.27 | 0.05 | −6.09 | 0.00 |
| Number of observations | 1,222 |  | 12,368 |  |  |  |  |
| Number of census tracts | 1,217 |  | 11,810 |  |  |  |  |

*Note:* The table reports the mean and standard deviation of different demographic variables for (i) individuals who completed the survey and (ii) individuals who did not complete the survey. Demographic characteristics are proxied by Census tract data. The last two columns report t-statistics and p-values of a two-sided t-test on the equality of means. Census tract socio-demographic data downloaded from https://data2.nhgis.org/main.

**Table A.6:** Subjective Risk Beliefs and Risk Exposure

|  | Movie | Restaurant | Transport | Service | Grocery | Severity index |
| --- | --- | --- | --- | --- | --- | --- |
| *Panel A. Correlation with demographics* |  |  |  |  |  |  |
| Age 30 to 44 | −0.016<br>(0.040) | −0.037<br>(0.039) | −0.059<br>(0.036) | −0.000<br>(0.036) | −0.023<br>(0.038) | −0.014<br>(0.026) |
| Age 45 to 59 | −0.014<br>(0.036) | −0.056<br>(0.037) | −0.047<br>(0.033) | −0.018<br>(0.036) | −0.045<br>(0.037) | 0.008<br>(0.027) |
| Age 60+ | −0.002<br>(0.036) | −0.050<br>(0.037) | −0.039<br>(0.036) | −0.010<br>(0.035) | −0.091***<br>(0.034) | 0.089***<br>(0.027) |
| Male | −0.062**<br>(0.027) | −0.077***<br>(0.026) | −0.082***<br>(0.025) | −0.057**<br>(0.023) | −0.068***<br>(0.024) | −0.024<br>(0.015) |
| Non-Hispanic Black | 0.024<br>(0.052) | 0.066<br>(0.064) | 0.074<br>(0.047) | 0.063<br>(0.060) | 0.040<br>(0.056) | 0.124***<br>(0.023) |
| Hispanic | 0.109***<br>(0.040) | 0.090**<br>(0.040) | 0.088***<br>(0.033) | 0.091**<br>(0.043) | 0.088**<br>(0.039) | 0.085***<br>(0.027) |
| Married | 0.005<br>(0.025) | 0.006<br>(0.027) | −0.004<br>(0.027) | 0.003<br>(0.025) | −0.026<br>(0.028) | −0.029**<br>(0.014) |
| Bachelor's degree+ | −0.025<br>(0.027) | −0.028<br>(0.024) | −0.002<br>(0.025) | 0.004<br>(0.024) | −0.064***<br>(0.021) | −0.076***<br>(0.013) |
| Essential worker | 0.014<br>(0.026) | −0.010<br>(0.027) | 0.011<br>(0.026) | 0.003<br>(0.032) | 0.021<br>(0.031) | −0.024<br>(0.015) |
| Has health conditions | 0.056**<br>(0.024) | 0.051**<br>(0.023) | 0.089***<br>(0.023) | 0.067***<br>(0.023) | 0.042*<br>(0.025) | 0.070***<br>(0.015) |
| Mean of Dep. Var. | 0.56 | 0.52 | 0.62 | 0.53 | 0.40 | 0.44 |
| Std. Dev. of Dep. Var. | 0.29 | 0.28 | 0.28 | 0.28 | 0.26 | 0.19 |
| No. of Obs. | 1,065 | 1,069 | 1,065 | 1,068 | 1,066 | 1,074 |
| *Panel B. Correlation with COVID-19 prevalence* |  |  |  |  |  |  |
| County-level COVID-19 cases<br>(per 1000) on May 7 | 0.003<br>(0.003) | 0.005*<br>(0.003) | 0.002<br>(0.002) | 0.004<br>(0.003) | 0.001<br>(0.002) | 0.001<br>(0.002) |
| Mean of Dep. Var. | 0.55 | 0.51 | 0.62 | 0.52 | 0.40 | 0.43 |
| Std. Dev. of Dep. Var. | 0.29 | 0.28 | 0.28 | 0.28 | 0.26 | 0.20 |
| No. of Obs. | 1,113 | 1,120 | 1,115 | 1,119 | 1,117 | 1,124 |

Note: Table presents the results of 12 separate OLS regressions specified in Equations 1 and 2. The point estimates are reported graphically in Figure 2. The dependent variables are the subjective belief about the risk of contracting COVID-19 when performing different economic activities or the severity of the disease conditional on contracting the virus. The measure of severe infection is an unweighted average of beliefs about the probability of having the following outcomes conditional on contracting the virus: having symptoms, needing medical care, needing hospitalization, not receiving treatment when needed, and death. All regression control for the number of days since a SIP order had been enacted. The sample is limited to 1,127 individuals, or 92.23% of survey respondents, who self-reported that a SIP order was in effect during the time of the survey. Survey regression weights are used to account for nonresponse. Stanford errors using jackknife replicate weights are reported in parentheses. \* for $p < .1$ \*\* for $p < .05$ and \*\*\* for $p < .01$ .

**Table A.7:** Reduction in Economic Activity and Geographic Prevalence of COVID-19

|  | Movies |  | Restaurant |  | Transport |  | Services |  | Grocery |  |
| --- | --- | --- | --- | --- | --- | --- | --- | --- | --- | --- |
|  | Policy | No policy | Policy | No policy | Policy | No policy | Policy | No policy | Policy | No policy |
| County-level COVID-19 cases (per 1000) on May 7 | 0.006<br>(0.004) | 0.011***<br>(0.004) | 0.006*<br>(0.003) | 0.010**<br>(0.004) | 0.019***<br>(0.005) | 0.013***<br>(0.004) | 0.010**<br>(0.004) | 0.009**<br>(0.004) | 0.003<br>(0.005) | 0.011**<br>(0.004) |
| Mean of Dep. Var. | 0.58 | 0.27 | 0.79 | 0.32 | 0.38 | 0.21 | 0.71 | 0.27 | 0.40 | 0.23 |
| Std. Dev. of Dep. Var. | 0.49 | 0.44 | 0.41 | 0.47 | 0.49 | 0.41 | 0.46 | 0.44 | 0.49 | 0.42 |
| No. of Obs. | 1,118 | 1,115 | 1,120 | 1,118 | 1,127 | 1,120 | 1,120 | 1,122 | 1,118 | 1,122 |
| Prevalence elasticity | 0.03 | 0.14 | 0.03 | 0.11 | 0.17 | 0.21 | 0.05 | 0.11 | 0.03 | 0.16 |

Note: Table presents the results of 10 separate linear regressions. The dependent variable is an indicator of whether the respondent decreased the activity by a lot with a separate model for actual behavior and hypothetical behavior in the absence of SIP. The independent variable is the prevalence of COVID-19 cases in May 2020. The sample is limited to 1,127 individuals, or 92.23% of survey respondents, who self-reported that a SIP order was in effect during the time of the survey. Survey regression weights are used to account for nonresponse. Stanford errors using jackknife replicate weights are reported in parentheses. \* for $p < .1$ \*\* for $p < .05$ and \*\*\* for $p < .01$ .

## Footnotes

1. A large literature examines both the potential and limitations of measuring quantitative subjective expectations (for reviews, see Manski 2004; Hurd 2009; Delavande 2014). The patterns of beliefs that we observe in our data appear consistent with the patterns reported by other key papers, including respondent use of focal points, overestimation of low risks, and overestimation based on salience (Viscusi et al. 1998; Fischhoff and Bruine De Bruin 1999; Bruin et al. 2000; Hurd 2009). For more extensive consideration of the literature on belief elicitation and these issues in our data, please see the Appendix A.1.

2. CDC Covid Tracker.

3. We discuss the extent to which our findings on the relationship between demographics and perceived risk are consistent with recent research on this relationship during the COVID-19 pandemic in Section 3.2.

4. Our results are consistent with those of Heffetz and Ishai 2021 who document that individual risk perceptions are better predictors of behavior than case count beliefs.

5. The fact that individuals substantially overestimated the risk of COVID-19 may in itself lead to a reduction in externalities because individuals would over-invest in protective behavior even in the absence of government mandates. At the same time, recent evidence suggests that behavioral biases may work the other direction as well, generating a “fatalism effect” – if individuals believe that COVID is widespread and inevitable, they may instead not take precautions (Akesson et al. 2020). We thank an anonymous referee for highlighting this point.

6. Individuals’ knowledge of whether there was a SIP order in their county of residence was nearly always accurate. 1,100 out of 1,219 respondents correctly reported that SIP was in place in their place of residence (we verified the county’s SIP status as of April 27 2021 based on Lin 2020; Baker-McKenzie 2020; Mervosh et al. 2020; Semerad 2020; Sylte et al. 2020; Gross 2020). 22 individuals correctly reported that they were not subject to a SIP at the time of our survey. 70 individuals had an incorrect belief about having or not having a SIP order in their area, while 27 reported not knowing if they were subject to a SIP.

7. https://covid.cdc.gov/covid-data-tracker

8. Appendix Table A.5 reports the pattern of selection into survey completion based on observable characteristics at the Census tract level. Individuals who completed the survey are more likely to live in Census tracts with a lower share of Hispanic or non-Hispanic Black population, higher share of college-educated population, lower share of households with incomes below the poverty line, higher share of population with a disability, and higher share of home ownership.

9. These results are consistent with the findings in Akesson et al. 2020 and Fetzer et al. 2021, who also document that individuals overestimate risk of COVID-19 in a survey environment.

10. To put our estimated magnitudes of activity reduction in context, it is helpful to compare them to the estimates of activity reductions from observational data. The estimates that are conceptually closest to ours are are reported in Farboodi et al. 2021, who find that retail and recreation activity fell a median of 33 percentage points, transit station activity by 25 percentage points, and workplace activity by 28 percentage points prior to SIPs and then as SIP orders spread across the country, the first three categories fell by a further 10, 15, and 11 percentage points. These large aggregate magnitudes of activity reductions are consistent with a substantial share of individuals in our sample reporting that they reduced these activities by more than 50%.

11. Note that this result is not mechanically driven by the existence of a SIP order as only individuals exposed to a SIP are included in our analysis.

12. It is again helpful to compare our findings to the estimates of the relative contribution of SIPs versus private behaviors from observational data. Maloney and Taskin 2020 estimate about a 60 pp drop in “voluntary” restaurant reservations and that SIP policies only accounted for 8pp out of this decline. Glaeser et al. 2021 find similarly limited impacts of SIPs on restaurants. Goolsbee and Syverson 2021 also conclude that formal restrictions contributed little to the changes in behavior. The results in Brzezinski et al. 2020 and Gupta et al. 2020, who estimate the SIP-induced versus “voluntary” choice of staying at home or general mobility, respectively, are very close to ours, estimating a 50/50 effect of SIP versus private decisions. The estimates in Cronin and Evans 2020 attribute a higher share of changes to private decisions, estimating the contribution of SIP to foot traffic in most industries to be up to a quarter of total reduction, with the rest attributable to private, self-regulating behavior.

